# Steroid Treatment Balance According To The Lymphocyte / White Blood Cells Ratio In COVID-19 Patients, A Retrospective Cohort Analysis

**DOI:** 10.1101/2022.02.16.22271033

**Authors:** Ferhat Arslan, Ali Mert, Mehmet Bayram, Handan Ankaralı, Haluk Vahaboglu

## Abstract

**Objectives:** Progressive respiratory failure is the main cause of clinical worsening in Coronavirus disease (COVID-19) patients. The decision to intubate during the follow-up of COVID-19 patients is critical because of high mortality rates.

**Methods:** We analyzed the COVID-19 related intubation and in-hospital mortality risk factors of patients admitted to two tertiary hospitals.

**Results:** Of the 275 patients included in the study, 44 (16%) were intubated, while 30 of them were patients (53%, 30/56) who had previously received steroid therapy. In 23 patients (77%) who received steroid therapy and were intubated, antiviral therapy was started in the first 6 days and Lymphocyte / White blood cells (LYM/WBC) ratios were lower than 0.18. The LYM/WBC ratio was found to be less than 0.12 in 14 patients who were intubated but did not receive steroid treatment before. 30(11%) of the patients included in the study died. While the number of deaths among those who did not receive steroid treatment was 10 (5%), it was 20 (36%) among the patients who did. Among in these 20 patients, it was observed that all 9 people who started antiviral treatment before 3 days, who were over 57 years old and whose oxygen saturation result was moderate or severe, died.

**Conclusions:** We think that the use of steroids in early period may be detrimental in rapidly progressive patients with lymphopenia that may be an independent marker of immune dysregulation.

**Box-ED Section:** *What is already known on the study topic?:* Progressive respiratory failure is the main cause of clinical worsening in COVİD-19 patients. Corticosteroid treatment has remarkable favourable effect on the prognosis.

*What is the conflict on the issue? Has it importance for readers?:* Corticosteroids is immunosupresive drug that have also nonspesific antienflammatory effect. Clinicians must consider multiple parameters such as patient status, disease period and existence of bacterial superinfection when to start corticosteroid treatment at bedside.

*How is this study structured?:* We conducted a retrospective study to analyze risk factors COVID-19 related intubation two cohorts. This study included patients diagnosed with COVID-19 induced pneumonia from March 21 to Apr 23,2020, at two university hospitals located on distinct sides of Istanbul province in Turkey.

*What does this study tell us?:* We think that the use of steroids in early period may be detrimental in rapidly progressive patients with lymphopenia that may be an independent marker of immune dysregulation

## Introduction

Despite the vaccination campaigns all over the world, the coronavirus disease (COVID-19) pandemic due to the novel severe acute respiratory syndrome coronavirus 2 (SARS-CoV-2) continues to be a serious public health problem with its new variants^1^.

Progressive respiratory failure is the main cause of clinical worsening in COVID-19 patients. This clinical worsening is usually the result of viral pneumonia in the first week of the disease and immunological pneumonitis added in the second week^2^. In a large registry, it has been claimed that dexamethasone treatment reduces 28-day mortality, especially in COVID-19 cases who need oxygen support ^3^. Subsequent prospective meta-analysis studies presented findings that contributed to the widespread use of steroids in clinical settings at worldwide ^4^. However, a recent meta-analysis emphasized that especially high-dose steroid therapy may have a negative effect on viral clearance and may cause clinical deterioration as well^5^.

This clinical deterioration can be caused by parenchymal diffuse infiltration, pulmonary thromboembolism and cardiac involvement alone or together, which will cause severe deterioration of oxygen exchange. Decision of intubation during patient follow-up has critical importance in terms of prognosis. Despite mortality rates decline after implementing more rationale approaches in intensive care unites (ICUs), all-cause mortality rate is still seen nearly 30% in intubated COVID-19 cases ^6,7^. It is necessary to determine the clinical and treatment related risk factors for intubation, especially in patients apart from ICUs.

In this study, we emphasize the probable subgroup of covid-19 patients that may have not respond the corticosteroid treatment effectively in our retrospective cohort. Additionally, comparing clinical outcomes of various anti-viral regimens is of scientific interest in these patients’ group.

## Methods

### Study design

We conducted a retrospective study to analyze risk factors for COVID-19 related intubation two cohorts. This study included patients diagnosed with COVİD-19 induced pneumonia from March 21 to Apr, 23,2020, at two university hospitals located on distinct side of the Istanbul province in Turkey.

Turkish Health Ministry had been recommended hydroxychloroquine (HCQ) plus azithromycin or favipiravir (FVP) in the treatment of COVID-19 patients in the first surge. While our center exclusively uses lopinavir-ritonavir (LPV/r) plus doxycycline (Dox) in in the management of moderate to severe COVID-19 cases, the other center had applied the recommendations of scientific committee of the government.

The ethics committee of the Istanbul Medipol University approved this study (No: 10840098-604.01.01-E.14713). Ethics committee waived obtaining informed consent.

### Centers, Patients and Definitions

We hereafter will describe centers according to the treatment protocol instead of the center name. Specifically, the center administering LPV/r plus Dox is referred to as “Center 1,” and that administering a range of treatment protocols approved by the Health Ministry is referred to as “Center 2.”

Cases were patients >18 years old who were hospitalized and found positive for the novel SARS-CoV-2 RNA following nasopharyngeal swab testing using reverse transcription polymerase chain reaction (RT-PCR). The LPV/r plus Dox cohort consisted of moderate to severe patients, since mild patients were home isolated with a different treatment protocol.

It was determined that a mild case had no respiratory symptoms, a moderate case had any signs of respiratory dysfunction, and a severe case was acute respiratory failure and required intensive care unit support by invasive or non-invasive means.

Study covariates included age (years), gender, lymphocyte count, white blood cell count, body temperature (°C) at admission, O_2_ saturation (%) at admission, respiration rate per minute at admission, the elapsed time between the onset of symptoms and hospitalization, history of hypertension, and Angiotensin-converting enzyme (ACE) inhibitors usage. The study outcome measures are described below. We extracted all data from medical records.

FVP 600 mg (10 days), HCQ 200 mg (10 days), LPV/r 200 mg and Dox 100 mg (7 days), all were administered twice daily via the oral route. Azithromycin was administered 250 mg (5 days) perioral once daily. For FVP, HCQ and azithromycin loading doses were given on the first day.

We analyzed the need for intubation as the primary outcome and in-hospital mortality as a secondary outcome. The need for intubation seems to be a more sensitive measure to compare the efficacy of the two different antiviral drugs and steroids in this settings. Mortality is not a reliable primary outcome especially after ICU admission in Covid-19 patients due to several ICU related events. Therefore, we give mortality prediction CT as supplementary files.

### Statistical Analysis

Descriptive statistics of the measurements were calculated as mean, standard deviation, and count and percent frequencies depending on the types of variables. In the study, two outcome measures were evaluated, namely the intubation status and the death status of the patients. First of all, the relationships between each outcome and demographic, clinical, and laboratory findings were examined with univariate tests as Pearson chi-square test and independent samples t-test. The results of these analyzes were used as preliminary information in the multivariate models established to examine risk factors. After univariate tests, risk factors were taken together in multivariate models and their adjusted effects on primary and secondary outcome variables were evaluated.

In this evaluation, multiple binary logistic regression model (LR), Support Vector Machine (SVM), Classification Tree (CT), and Random Forest (RF) algorithms were used. 10-fold cross-validation method of estimating prediction error was used. For comparing performances of these models and algorithms in predicting intubation and ex status was used with the area under the curve (AUC), accuracy, F1 ratio, precision, and recall statistics. In addition, according to the logistic regression model, nomograms were created for the risk of intubation and ex status.

According to statistical tests results, P values are accepted as statistically significant if less than 0,05.

SPSS (ver. 23), WEKA (ver. 3.8.5), and R language and environment (R Core Team, 2021) were used for all statistical computations.

## Results

During the study period, 161 and 114patients were followed in the Group 1 cohort and the Group 2 cohort, respectively. **Table 1** and **Table 2** show univariate analysis of categorical and continue variables according to the intubation and 28 days mortality as primary and secondary outcomes, respectively. Statistically significant (P < 0.05) variables used in multivariate models.

**Table 1.** Categorical variables according to the intubation and mortality.

| 1. Risk Factors |  | Code | Intubation |  |  |  | P* | Status |  |  |  | P* |
| --- | --- | --- | --- | --- | --- | --- | --- | --- | --- | --- | --- | --- |
|  |  |  | No |  | Yes |  |  | Survive |  | Ex |  |  |
|  |  |  | n | % | n | % |  | n | % | n | % |  |
| Steroid | No | 0 | 205 | 93,6 | 14 | 6,4 | <0,001 | 209 | 95,4 | 10 | 4,6 | <0,001 |
|  | Yes | 1 | 26 | 46,4 | 30 | 53,6 |  | 36 | 64,3 | 20 | 35,7 |  |
| Center | Center 1 | 1 | 136 | 84,5 | 25 | 15,5 | 0,800 | 141 | 87,6 | 20 | 12,4 | 0,339 |
|  | Center 2 | 2 | 95 | 83,3 | 19 | 16,7 |  | 104 | 91,2 | 10 | 8,8 |  |
| Gender | Male | 1 | 125 | 83,9 | 24 | 16,1 | 0,958 | 134 | 89,9 | 15 | 10,1 | 0,626 |
|  | Female | 2 | 106 | 84,1 | 20 | 15,9 |  | 111 | 88,1 | 15 | 11,9 |  |
| O2 Saturation | Mild | 1 | 141 | 90,4 | 15 | 9,6 | <0,001 | 148 | 94,9 | 8 | 5,1 | <0,001 |
|  | Moderate | 2 | 64 | 87,7 | 9 | 12,3 |  | 68 | 93,2 | 5 | 6,8 |  |
|  | Severe | 3 | 25 | 56,8 | 19 | 43,2 |  | 28 | 63,6 | 16 | 36,4 |  |
| Favipravir | Center 1 | 0 | 136 | 84,5 | 25 | 15,5 | 0,883 | 141 | 87,6 | 20 | 12,4 | 0,464 |
|  | Yes | 1 | 40 | 81,6 | 9 | 18,4 |  | 46 | 93,9 | 3 | 6,1 |  |
|  | No | 2 | 55 | 84,6 | 10 | 15,4 |  | 58 | 89,2 | 7 | 10,8 |  |
| Azitromycin | Center 1 | 0 | 136 | 84,5 | 25 | 15,5 | 0,197 | 141 | 87,6 | 20 | 12,4 | 0,185 |
|  | Yes | 1 | 65 | 87,8 | 9 | 12,2 |  | 70 | 94,6 | 4 | 5,4 |  |
|  | No | 2 | 30 | 75,0 | 10 | 25,0 |  | 34 | 85,0 | 6 | 15,0 |  |
| Tocilizumab | Center 1 | 0 | 136 | 84,5 | 25 | 15,5 | 0,021 | 141 | 87,6 | 20 | 12,4 | 0,435 |
|  | Yes | 1 | 21 | 67,7 | 10 | 32,3 |  | 27 | 87,1 | 4 | 12,9 |  |
|  | No | 2 | 74 | 89,2 | 9 | 10,8 |  | 77 | 92,8 | 6 | 7,2 |  |
| Hidroxychloroquin | Center 1 | 0 | 136 | 84,5 | 25 | 15,5 | 0,007 | 141 | 87,6 | 20 | 12,4 | 0,010 |
|  | Yes | 1 | 80 | 88,9 | 10 | 11,1 |  | 86 | 95,6 | 4 | 4,4 |  |
|  | No | 2 | 15 | 62,5 | 9 | 37,5 |  | 18 | 75,0 | 6 | 25,0 |  |
| Hipertension | No | 0 | 153 | 87,9 | 21 | 12,1 | 0,018 | 164 | 94,3 | 10 | 5,7 | <0,001 |
|  | Yes | 1 | 77 | 77,0 | 23 | 23,0 |  | 80 | 80,0 | 20 | 20,0 |  |
| ACE inhibitors | No | 0 | 194 | 84,0 | 37 | 16,0 | 0,986 | 208 | 90,0 | 23 | 10,0 | 0,246 |
|  | Yes | 1 | 37 | 84,1 | 7 | 15,9 |  | 37 | 84,1 | 7 | 15,9 |  |
| ATIT | 0-3 day | 1 | 41 | 69,5 | 18 | 30,5 | 0,002 | 45 | 76,3 | 14 | 23,7 | 0,002 |
|  | 4-6 day | 2 | 84 | 85,7 | 14 | 14,3 |  | 91 | 92,9 | 7 | 7,1 |  |
|  | >=7 day | 3 | 106 | 89,8 | 12 | 10,2 |  | 109 | 92,4 | 9 | 7,6 |  |

ATIT: Antiviral treatment initiation time (after symptoms started)

**Table 2.** Categorical variables according to the intubation and mortality

|  | Intubation |  |  |  |  |  |  | Mortality |  |  |  |  |  |  |
| --- | --- | --- | --- | --- | --- | --- | --- | --- | --- | --- | --- | --- | --- | --- |
|  | No |  |  | Yes |  |  | P* | Survived |  |  | Died |  |  | P* |
|  | N | Mean | SD | N | Mean | SD |  | N | Mean | SD | N | Mean | SD |  |
| Age | 231 | 56,95 | 15,94 | 44 | 67,16 | 14,57 | **<0,001** | 245 | 56,84 | 15,68 | 30 | 72,87 | 12,57 | <0,001 |
| Lymphocyte | 228 | 1,25 | ,52 | 44 | 1,25 | 1,00 | 0,984 | 242 | 1,25 | ,53 | 30 | 1,23 | 1,12 | 0,928 |
| Neutrophil | 133 | 4,66 | 3,51 | 25 | 7,58 | 5,37 | **0,014** | 138 | 4,88 | 3,96 | 20 | 6,82 | 3,85 | 0,041 |
| WBC | 228 | 6,23 | 3,26 | 44 | 9,60 | 5,09 | **<0,001** | 242 | 6,46 | 3,65 | 30 | 9,31 | 4,20 | 0,001 |
| Fever | 230 | 34,17 | 10,40 | 42 | 34,88 | 9,56 | 0,681 | 244 | 34,06 | 10,56 | 28 | 36,20 | 6,96 | 0,156 |
| Respiratory rate | 222 | 21,16 | 4,30 | 41 | 23,71 | 6,55 | **0,021** | 236 | 21,42 | 4,53 | 27 | 22,74 | 6,71 | 0,328 |
| LYM/<br>WBC ratio | 228 | ,23 | ,10 | 44 | ,15 | ,11 | **<0,001** | 242 | ,22 | ,10 | 30 | ,14 | ,11 | <0,001 |

WBC: White blood cells, LYM: Lymphocyte

The factors presented in **Table 3** were determined as risk factors according to univariate tests (P<0.05). A total of 9 risk factors were found associated with intubation of the patient, and 7 risk factors were found associated with mortality.

**Table 3.** Risk factors included in multivariate models to investigate their relationship with intubation and mortality

| Intubation (0=No, 1=Yes) | Mortality (0=Survived, 1=Died) |
| --- | --- |
| Steroid treatment (0=No, 1=Yes) | Steroid treatment (0=No, 1=Yes) |
| PaO2 (1=Mild, 2=Moderate, 3=Severe) | PaO2 (1=Mild, 2=Moderate, 3=Severe) |
| Tocilizumab (0=Kaletra, 1=Tosilizumab, 2=No) | --- |
| HCQ (0=Kaletra, 1=HCQ, 2=No) | HCQ (0=Kaletra, 1=HCQ, 2=No) |
| HT (0=No, 1=Yes) | HT (0=No, 1=Yes) |
| ATIT (1; 0-3 day, 2; 4-6 day, 3; 7 day and more) | ATIT (1;0-3 day, 2;4-6 day, 3;7 day and more) |
| Age | Age |
| Respiratory rate | --- |
| LYM /WBC ratio | LYM /WBC ratio |

We did multiple logistic regression model (LR), support vector machines (*SVMs*), Classification Tree (CT) and Random Forest (RF) methods in predicting intubation status. The lowest error in classifying non-intubated individuals belongs to the SVM model, and the lowest error in estimating intubated individuals belongs to the CT model. The fact that the number of intubated patients is 44 in total may adversely affect the classification success. The overall classification success (Accuracy) of the four algorithms were similar.

The nomogram for the intubation risk obtained according to the logistic regression model is given in **Figure 1**. The risk of intubation increases as a person’s risk score increases. The most contributing risk factors to this score were steroid treatment, increased respiratory rate, initiation of antiviral treatment between 4-6 days, receiving HCQ and Tocilizumab treatment even with a low contribution or no treatment, low LYM/WBC ratio, advanced age, severe O2 saturation, and presence of HT.

**Figure 1.**
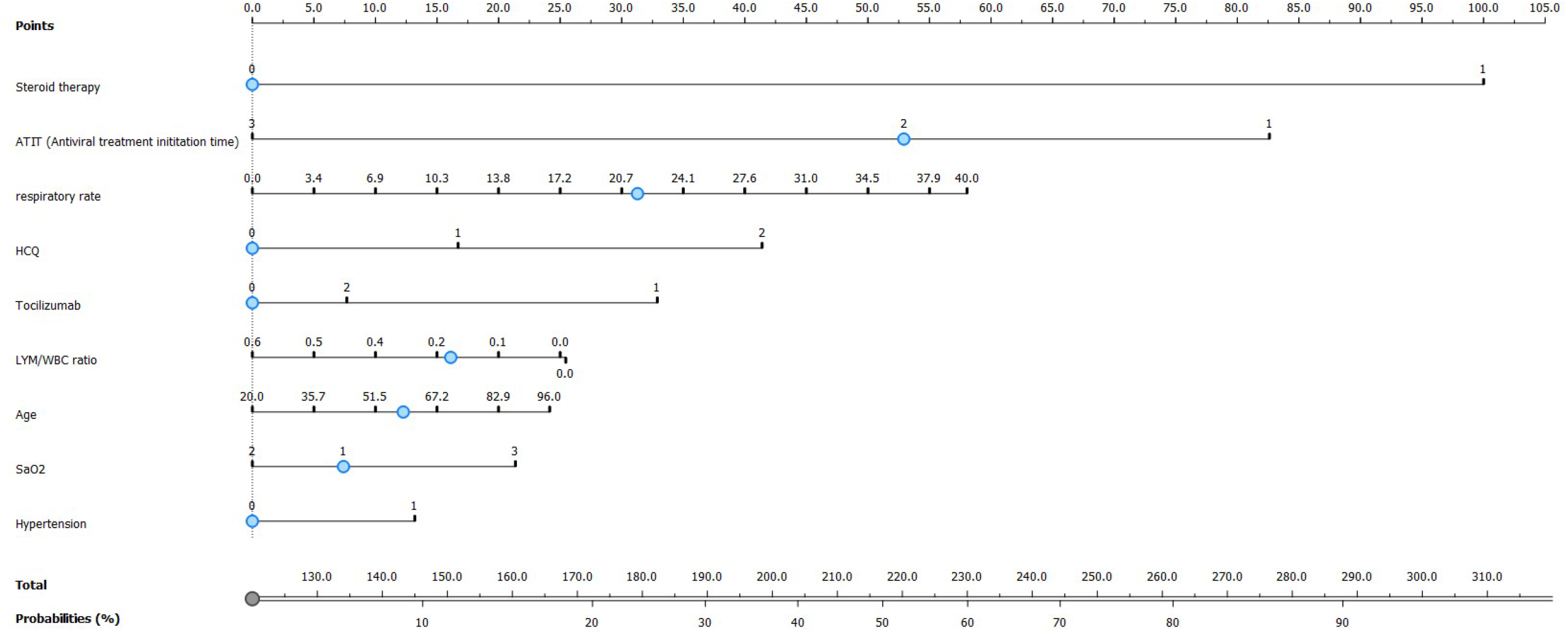
The nomogram for the intubation risk

Classification tree results for intubation is given in **Figure 2**. Receiving steroid therapy was the most significant risk factor for being intubated. Of the 275 patients included in the study, 44 (16%) were intubated, while 30 of them were patients (53%, 30/56) who had previously received steroid therapy. None of the antiviral regimes have impact on the prognosis that make decision point in CT.

**Figure 2.**
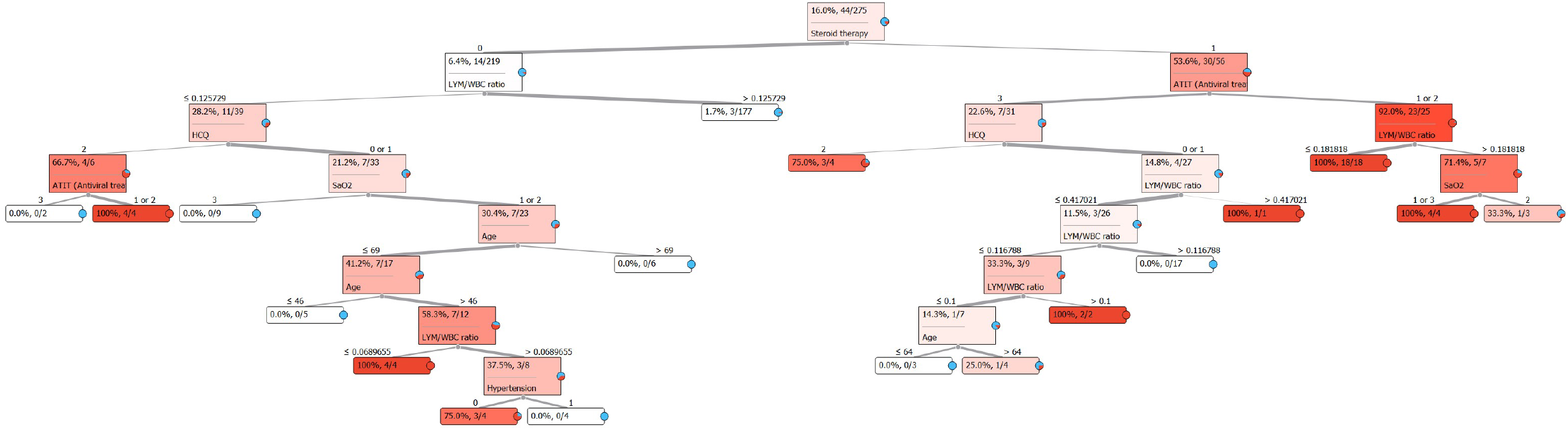
Classification Tree for intubation

In 23 patients (77%) who received steroid therapy and were intubated, antiviral therapy was started in the first 6 days and LYM/WBC ratios were lower than 0.18. The LYM/WBC ratio was found to be less than 0.12 in 14 patients who were intubated but did not receive steroid treatment before. Decision points in the CT that a very small number of patients participated do not have the ability to indicate a spesific trend.

30 (11%) of the patients included in the study died. While the number of deaths among those who did not receive steroid treatment was 10 (5%), it was 20 (36%) among the patients who did. Among in these 20 patients, it was observed that all 9 people who started antiviral treatment before 3 days, who were over 57 years old and whose oxygen saturation result was moderate or severe, died. In other cases who received steroid treatment and died, being over 68 years seems to be a relatively important factor. According to the CT method, steroid use, age (>68), low oxygen saturation and LYM/WBC ratio (<0.09) are seen as risk factors for mortality. (**s. Figure 1**. Classification Tree for mortality risk).

## Discussion

This retrospective clinical study confers patients who were followed up in the first surge of the COVID-19 pandemic, with low lymphocyte ratio and steroid intervention in early period at admission had poor prognosis. Other observational and randomized clinical studies support this finding as well^8^. The lack of widespread use of steroid therapy in early-stage COVID-19 patients may provide data on the outcomes of steroid administration in those with and without severe lymphopenia at baseline. In our study, it is noteworthy that the risk of intubation was increased in patients with severe lymphopenia and steroid treatment in early period. The definition of lymphopenia varies in studies^9^. Therefore, we thought that it would be a more accurate approach to evaluate the dynamic variability of the bone marrow with a ratio. While the risk of intubation increased significantly in those with severe lymphopenia (LYM/WBC ratio <0.06) in steroid non-received group, this rate increased to 0.18 in those who started antiviral therapy within 7 days of symptoms started, in steroid received group. Despite early initiation of antiviral therapy, clinical rapid progression may be thought to be mediated by specific immune dysregulation in some patients^10^. It is not possible to define these and similar immune deficiencies in clinical practice. Here, we would like to draw attention to steroid therapy which is widely used in COVID-19 disease, may affect prognosis negatively in some patients with lymphopenia.

Lu et al. claimed in their observational studies that steroid treatment may contribute positively to the prognosis in patients with lymphopenia ^11^ Interestingly, in the same study, it was emphasized that early and high-dose steroid therapy is poor prognostic. In their observational clinical studies, Cai et al, found that steroid treatment have no positive effect on COVID-19 prognosis in patients with a lymphocyte-neutrophil ratio less than 6.11 ^12^. In particular, CD8+ T cells have been shown to be an independent variable in the immune response and recovery of COVID-19^13^. The reducing effect of prednisone on the number of peripheral T cells has been known for many years^14^. Clinicians may use high-dose and long-term steroids to suppress severe inflammation, which they define as cytokine storm, in rapidly progressing cases especially presented with acute respiratory distress syndrome (ARDS). If patients with rapidly progressing COVID-19 also have severe lymphopenia, the limited effect of steroid therapy on prognosis may not be as expected in ARDS due to other causes^15^.

## Limitations

Our study has limitations, including its retrospective nature and some violations of comparability at the two hospital settings. The compliance of outpatient treatments could not be followed up in patients discharged early. Unrecorded data about the patients who remained hospitalized, especially at the ICU, after the study was complete may cause the results to be biased. The adverse effect of steroids on prognosis may be an overestimated variable, especially in rapidly progressing patients.

## Conclusion

We think that the use of steroids in early period may be detrimental in rapidly progressive patients with lymphopenia that may be an independent marker of immune dysregulation.

## Data Availability

All data produced in the present study are available upon reasonable request to the authors

## Acknowledgement

We are thankfull to study group (Sacit Icten, Pinar Ergen, Ozlem Aydin, Ayse Canan Ucisik, Fatma Yilmaz-Karadag, Hulya Caskurlu, Bahadır Ceylan, Selda Aydın, Muhammed Emin Akkoyunlu, Mustafa Düger, Okan Derin, Abdullah Kansu, Sedef Başgönül, Tulin Akarsu-Ayazoglu, Hasan Kocoglu, Yasemin Cag) for data acquisition.

## Figures

Supp.Figure 1. Classification Tree for mortality risk

