## Supplementary figures and images for "Steroid Treatment Balance According To The Lymphocyte / White Blood Cells Ratio In COVID-19 Patients, A Retrospective Cohort Analysis"

### s.Figure 1

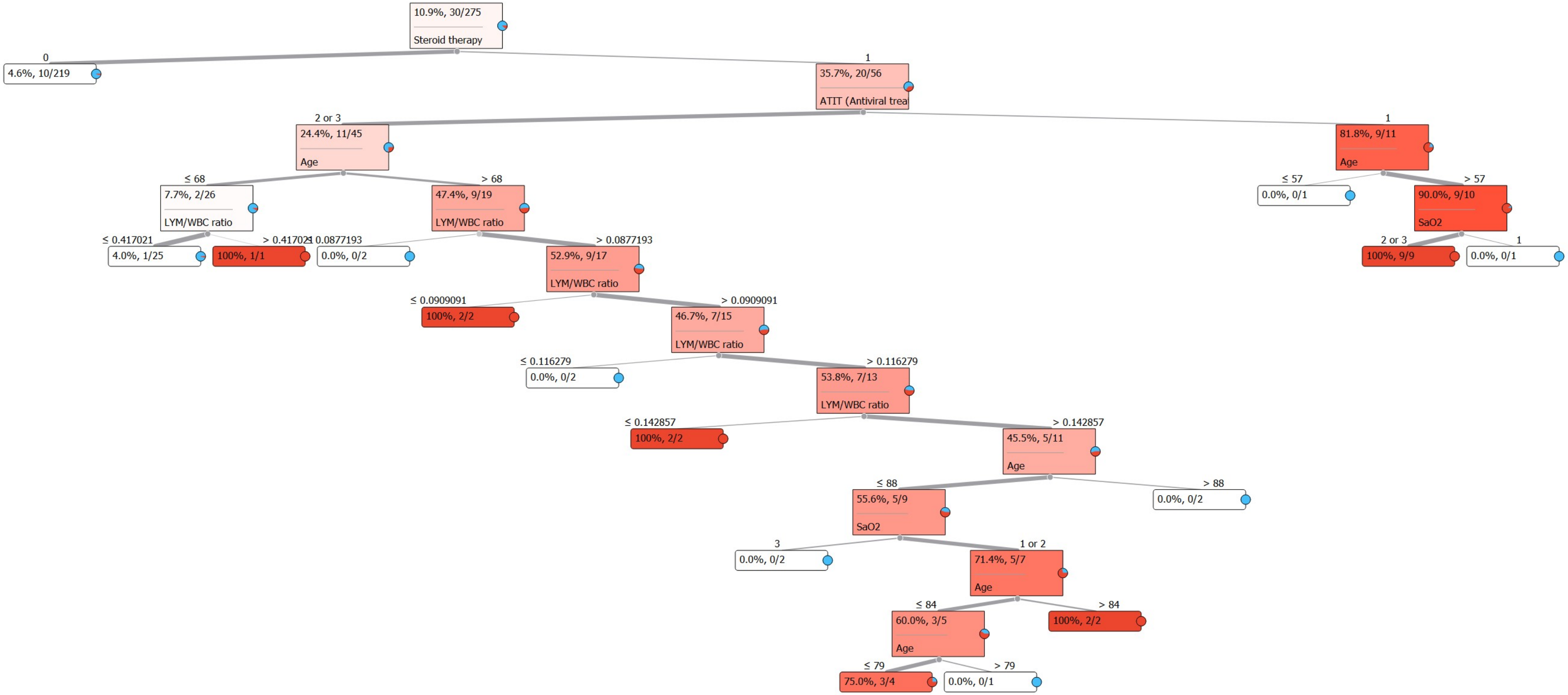
